# MENSA, a Media Enriched with Newly Synthesized Antibodies, to Identify SARS-CoV-2 Persistence and Latent Viral Reactivation in Long-COVID

**DOI:** 10.1101/2024.07.05.24310017

**Authors:** Natalie S. Haddad, Andrea Morrison-Porter, Hannah Quehl, Violeta Capric, Pedro A. Lamothe, Fabliha Anam, Martin C. Runnstrom, Alex D. Truong, Adviteeya N. Dixit, Matthew C. Woodruff, Anting Chen, Jiwon Park, Doan C. Nguyen, Ian Hentenaar, Caroline Y. Kim, Shuya Kyu, Brandon Stewart, Elizabeth Wagman, Hannah Geoffroy, Daniel Sanz, Kevin S. Cashman, Richard P. Ramonell, Monica Cabrera-Mora, David N. Alter, John D. Roback, Michael C. Horwath, James B. O’Keefe, Alexandra W. Dretler, Ria Gripaldo, Samantha M. Yeligar, Ted Natoli, Viktoria Betin, Rahulkumar Patel, Kennedy Vela, Mindy Rodriguez Hernandez, Sabeena Usman, John Varghese, Anum Jalal, Saeyun Lee, Sang N. Le, R. Toby Amoss, John L. Daiss, Ignacio Sanz, F. Eun-Hyung Lee

## Abstract

Post-acute sequelae of SARS-CoV-2 (SARS2) infection (PASC) is a heterogeneous condition, but the main viral drivers are unknown. Here, we use MENSA, Media Enriched with Newly Synthesized Antibodies, secreted exclusively from circulating human plasmablasts, to provide an immune snapshot that defines the underlying viral triggers. We provide proof-of-concept testing that the MENSA technology can capture the new host immune response to accurately diagnose acute primary and breakthrough infections when known SARS2 virus or proteins are present. It is also positive after vaccination when spike proteins elicit an acute immune response. Applying the same principles for long-COVID patients, MENSA is positive for SARS2 in 40% of PASC vs none of the COVID recovered (CR) patients without any sequelae demonstrating ongoing SARS2 viral inflammation only in PASC. Additionally, in PASC patients, MENSAs are also positive for Epstein-Barr Virus (EBV) in 37%, Human Cytomegalovirus (CMV) in 23%, and herpes simplex virus 2 (HSV2) in 15% compared to 17%, 4%, and 4% in CR controls respectively. Combined, a total of 60% of PASC patients have a positive MENSA for SARS2, EBV, CMV, and/or HSV2. MENSA offers a unique antibody snapshot to reveal the underlying viral drivers in long-COVID thus demonstrating the persistence of SARS2 and reactivation of viral herpes in 60% of PASC patients.

**Graphical abstract:** 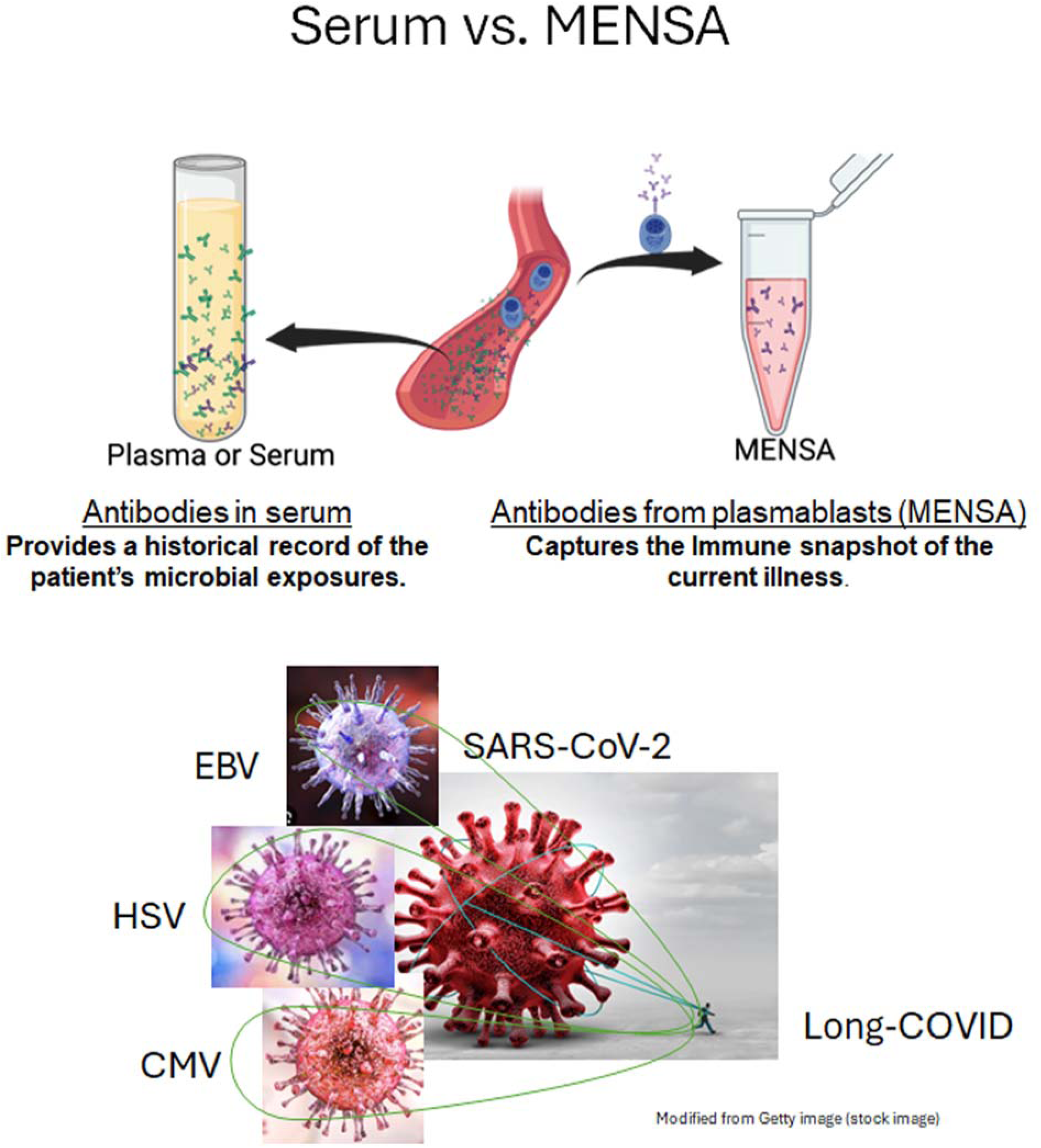

## INTRODUCTION

In December 2019, the world was changed when the SARS-CoV-2 (SARS2) virus was identified in Wuhan, China and rapidly spread throughout the world. The first U.S. case was identified in January 2020^1^, and by March 2020 the World Health Organization (WHO) had officially declared COVID-19 to be a global pandemic. By the end of 2020, there had been a total of 79 million reported cases and over 1.7 million deaths globally^2^. Vaccines and antiviral therapies allowed us to combat this new global threat and emerge from this devastating pandemic^3,4^. However, after many in the US received the primary mRNA vaccines, a new Delta virus surged, followed by the Omicron (B.1.1.529) variant by the end of 2021, which had increased transmissibility^5^.

In addition to new viral variant infections, some patients suffered from sequelae after the initial acute infection. Long-COVID or post-acute sequelae of SARS-CoV-2 infection (PASC) is a condition described as ongoing, relapsing, or new symptoms present after the acute phase of the infection. Incidence of PASC was notable in approximately 10% of patients after acute infection^6^. Definitions by the CDC used symptoms > 30 days after acute infection whereas the WHO described continuation or development of new symptoms 3 months after the initial SARS2 infection, with symptoms lasting for at least 2 months with no other explanation^7^. The diversity of symptoms and differences in plasma proteomics between inflammatory and quiescent PASC subsets attest to the heterogeneity of long-COVID^8^. Additionally, many multiomic studies have identified metabolic and inflammatory derangements such as decreased cortisol or serotonin levels^9–12^, complement dysregulation^13^, and alternations of cytotoxic T cell, atypical B cell, or neutrophil signatures in PASC patients compared to adults who recovered from SARS2 without sequelae^9,10^. Some studies suggest that these inflammatory changes may be triggered by viral persistence of SARS2 and/or reactivation of latent EBV^10,14^. Interestingly, these studies used viral PCR testing which is known to be less sensitive in the blood or requires ultrasensitive spike antigen tests. Another study showed higher serum levels of EBV antibodies in PASC patients (47%) compared to healthy adults (28%) but there was significant overlap between the two groups^9,15^ due to serum antibodies in response to a previous infection, confounding the observation of new or ongoing immune responses.

In this study, we use a novel diagnostic platform whereby we capture antibodies secreted from plasmablasts or newly-minted antibody secreted cells (ASC). This method can identify new or repeat infections despite elevated serum antibodies since these ASC appear in circulation shortly after infection or vaccination, then rapidly disappear from the blood^16–20^ ^21,22^. By capturing antibodies from these special ASC in a new matrix called <u>M</u>edia <u>E</u>nriched with <u>N</u>ewly <u>S</u>ynthesized <u>A</u>ntibodies (MENSA), we provide a signature response from only the new illness.

MENSA antibodies often appear prior to seroconversion and differ from serum antibodies, which confound results with the patient’s entire historical microbial record. In prior studies, we have successfully used MENSA to diagnose acute Lyme disease, *Clostridioides difficile*, *Streptococcus pneumoniae*, and deep bone/tissue infections with *Staphylococcus aureus* ^19,23–27^. In all, the novel MENSA assay can successfully diagnose repeat bacterial and viral infections from a single blood sample even when serum antibody titers are extremely high, showing the assay’s exceptional ability to resolve complexity of antibody signals.

Here, we provide proof-of-concept testing that the MENSA technology can capture the new host immune response to accurately diagnose acute primary and breakthrough infections when known SARS2 virus or proteins are present. It is also positive after vaccination when spike proteins elicit an acute immune response. Applying the same principles for long-COVID patients, we use MENSA to identify SARS2, EBV, CMV, and/or HSV2 as the underlying viral drivers in 60% of PASC patients. With a single blood sample, this novel diagnostic assay shows persistence of SARS2 and/or reactivation of latent herpes viruses in long-COVID patients.

## RESULTS

### Patient enrollment

For the purpose of measuring serum and MENSA responses against SARS2 during infection and after vaccination, we enrolled a total of 241 adults between 2020 and 2024 at Emory University in Atlanta, GA and collected blood to generate the MENSA matrix and serum samples (**Table 1**). During the first year of the pandemic in 2020, we enrolled 110 adults with PCR-positive nasopharyngeal swabs (NPS) during their primary SARS2 infection. Fifty-four adults were outpatients with mild/moderate (M/M) disease as defined by the NIH criteria^28^, and 56 adults had severe/critical (S/C) illness in the Intensive Care Units. From the 54 M/M adults, we collected 59 blood samples: 16 during acute infection (within 30 days post-symptom onset (DPSO)) and 43 during convalescence (60 DPSO). Five patients provided both acute and convalescent samples. Of the 56 S/C patients, we had 60 samples: 40 provided acute illness time points and 20 at convalescence. Four patients provided samples at both time points. These patients will be referred to as the primary SARS2 infection population. We also enrolled 60 healthy adults who had no known exposure to SARS2 and provided blood samples during the initial lockdown period, between March and June 2020. This group is referred to as the healthy controls without prior SARS2 exposure.

**Table 1.** Demographics of Controls, COVID-19 patients, and Vaccinated Subjects.

| Subjects | Healthy Adults | SARS2 Infection |  | Vaccinated | Post-SARS Infection |  |
| --- | --- | --- | --- | --- | --- | --- |
|  |  | Mild/Moderate | Severe/Critical | Pfizer & Moderna | PASC | COVID Recovered |
|  | N = 60 | N = 54 | N = 56 | N = 11 | N = 61 | N = 25 |
| **Age**, years |  |  |  |  |  |  |
| (Mean ± SD) | (30 ± 9) | (40 ± 14) | (56 ± 14) | (40 ± 16) | (49 ± 12) | (41 ± 15) |
| **Sex**, N (%) |  |  |  |  |  |  |
| Males | 31 (52) | 18 (33) | 30 (54) | 2 (18) | 15 (25) | 7 (28) |
| Females | 29 (48) | 36 (67) | 26 (46) | 9 (82) | 46 (75) | 18 (72) |
| **Race**, N (%) |  |  |  |  |  |  |
| Caucasian | 41 (68) | 30 (56) | 17 (30) | 3 (27) | 24 (39) | 11 (44) |
| African American | 4 (7) | 17 (31) | 35 (63) | 3 (27) | 34 (56) | 9 (36) |
| Asian | 10 (17) | 4 (7) | 4 (7) | 5 (45) | 2 (3) | 3 (12) |
| Other | 5 (8) | 3 (6) | 0 (0) | 0 (0) | 1 (2) | 2 (8) |

Serum and MENSA samples were collected from 11 vaccinated subjects who had no SARS2 infection prior to their primary two-dose mRNA vaccine series (Pfizer n=5, Moderna n=6). We also enrolled 3 adults with Omicron breakthrough infections from December 2021-June 2022. Finally, we enrolled 61 PASC patients from the Emory Long-COVID clinic from January 2021 to February 2024 and compared against 25 COVID recovered patients without any sequelae (CR). Of note, some of the same subjects were enrolled in different groups. For example, some of the healthy controls were later vaccinated and/or tested positive for breakthrough infections in the 4 years of the study; some patients after acute primary and/or breakthrough COVID infections without any sequelae were also included in the CR groups.

### SARS2 RBD and N antigen selection

SARS2 antigens were selected for measuring responses to SARS2 infection and vaccination. Fluorescent bead assays were used as previously described for the SARS2 antigens with spike S1, S1 receptor binding domain (RBD), S1 N-terminal domain (NTD), S2, nucleocapsid (N), and ORF-3a ^29^. We measured the acute IgG antibody responses against all six antigens in 56 adults with primary acute SARS2 infection between 6-28 DPSO and compared them against 60 healthy control adults with no known SARS2 exposure. MENSA and serum antibody reactivity was significantly higher in the infected groups than in the healthy control group for each of the six antigens (**Supplementary figure 1**). Receiver operating characteristic (ROC) curves yielded Area Under the Curve (AUC) values of ≥0.89 for the four spike-associated proteins, with MENSA anti-RBD demonstrating the highest value, AUC=1.0 (**Supplementary figure 2**). To distinguish natural infection responses from vaccination responses, we examined the diagnostic potential of two non-spike proteins, N and ORF3a. N yielded much higher signals in the infected groups and a slightly higher AUC value than ORF3a. Therefore, we focus on anti-RBD and anti-N IgG for all subsequent analyses.

### Primary SARS2 infection responses in MENSA and serum

Primary infected patients were further divided based on the severity of their acute infections. During the acute stage of primary SARS2 infection, we observe a rise in both MENSA and serum anti-RBD IgG for M/M (69%, 88%) and S/C (95%, 95%) patients (**Fig. 1A,B**). We see a similar rise in MENSA and serum anti-N IgG for M/M (69%, 88%) and S/C (80%, 95%) patients as well (**Fig. 1C,D**). During convalescence (60-360 DPSO, 2-12 months after), MENSA levels drop significantly while serum levels rise quickly and remain elevated for months to years especially for anti-RBD IgG (**Fig. 1**). The MENSA levels from S/C were higher than in M/M patients during the acute infection because of higher frequencies of circulating early-minted ASC as previously shown by flow cytometry^30^. Additionally, for S/C compared to MM patients, serum levels rise higher during the acute illness and remain elevated during convalescence, suggesting more ASC survived in other tissues such as the spleen, bone marrow, or mucosal sites. During convalescence, 87% of MENSAs become negative, but are overall slightly higher than pre-pandemic levels. In all, MENSA rises during acute infection and then rapidly falls to negative whereas serum titers rise and remain elevated.

**Figure 1.**
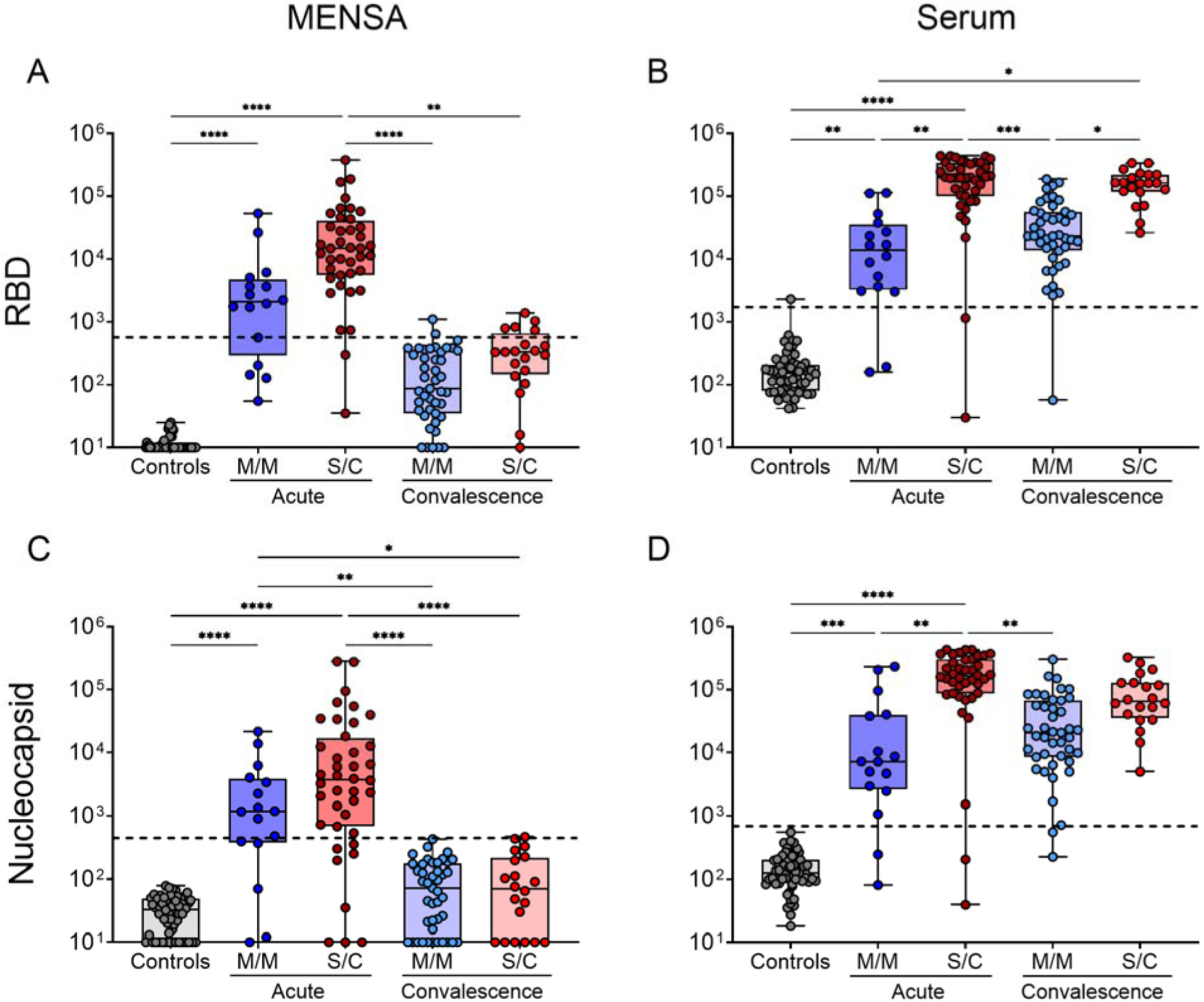
Primary wild type COVID-19 infection responses in MENSA and serum. Dot plots show IgG antibody reactivity against S1-RBD in the MENSA (A) and serum (B) of patients experiencing Wild Type SAR-CoV-2 infections in 2020. Similar results are also shown for anti-Nucleocapsid IgG in the MENSA (C) and serum (D). Samples from patients with acute Mild/Moderate (59 samples from 54 patients; blue dots) and Severe/Critical (60 samples from 56 patients; red dots) infections were collected less than 30 DPSO (acute) and/or after 60 DPSO (convalescence). Early pandemic healthy controls, with no prior exposure to SARS2 (n=60), are shown as black dots on the left of each panel. All units are represented as Median Fluorescent Intensity minus background (Net MFI). Dashed lines indicate the C_0_ threshold of positivity for each sample type and antigen. Serum C_0_s were calculated as the average Net MFI plus five standard deviations of the 60 healthy controls (RBD: 1724; N: 682). For MENSA C0s, a subset of the convalescent patients was identified as a confirmed COVID Recovered (CR) population (no sequelae; N=19) and was used as a contemporary control group to calculate the average Net MFI plus 3 standard deviations (RBD: 570; N: 441). Pair-wise comparisons were performed using the Kruskal-Wallis test in GraphPad Prism (unpaired, nonparametric test; ns p > 0.05, * p ≤ 0.05, ** p ≤ 0.01, *** p ≤ 0.001, **** p ≤ 0.0001).

### Determining C_0_ thresholds for positivity

Since the convalescent baseline MENSA negative values could be slightly higher than in pre-pandemic controls, we identify a subset of the convalescent patients from Fig. 1 as COVID Recovered (CR) who had fully recovered with no sequelae (N=19). CR MENSA samples were used as controls to calculate the MENSA C_0_ using the average Net MFI plus 3 standard deviations. In contrast to MENSA, serum levels rise and remain high indefinitely in both CR and PASC patients; therefore, the serum C_0_ values are calculated using the average Net MFI plus 5 standard deviations of the 60 healthy controls prior to SARS2 exposure. These calculated Net MFI C_0_ values for MENSA (RBD: 570; N: 441) and serum (RBD: 1724; N: 682) are used to distinguish positive and negative samples in Figures 1-3, 5, and Supplementary figures 3, 4.

### MENSA and serum from primary and booster mRNA vaccination

Similar to observations in primary acute SARS2 infections, adults receiving the COVID-19 vaccination, with no known prior exposure, have positive MENSA IgG for SARS2 but only to RBD and not specific to N since only spike proteins are engineered in the mRNA vaccines (**Fig. 2**). This rise in MENSA IgG to RBD 1-2 weeks after the first dose further increases to a higher peak after the second dose. MENSA anti-RBD antibodies decline to negative levels prior to the third vaccine dose and then increase again within a week after the third booster (**Fig. 2A**). Serum anti-RBD antibody levels increase after dose one and remain elevated throughout months to years (**Fig. 2B**).

**Figure 2.**
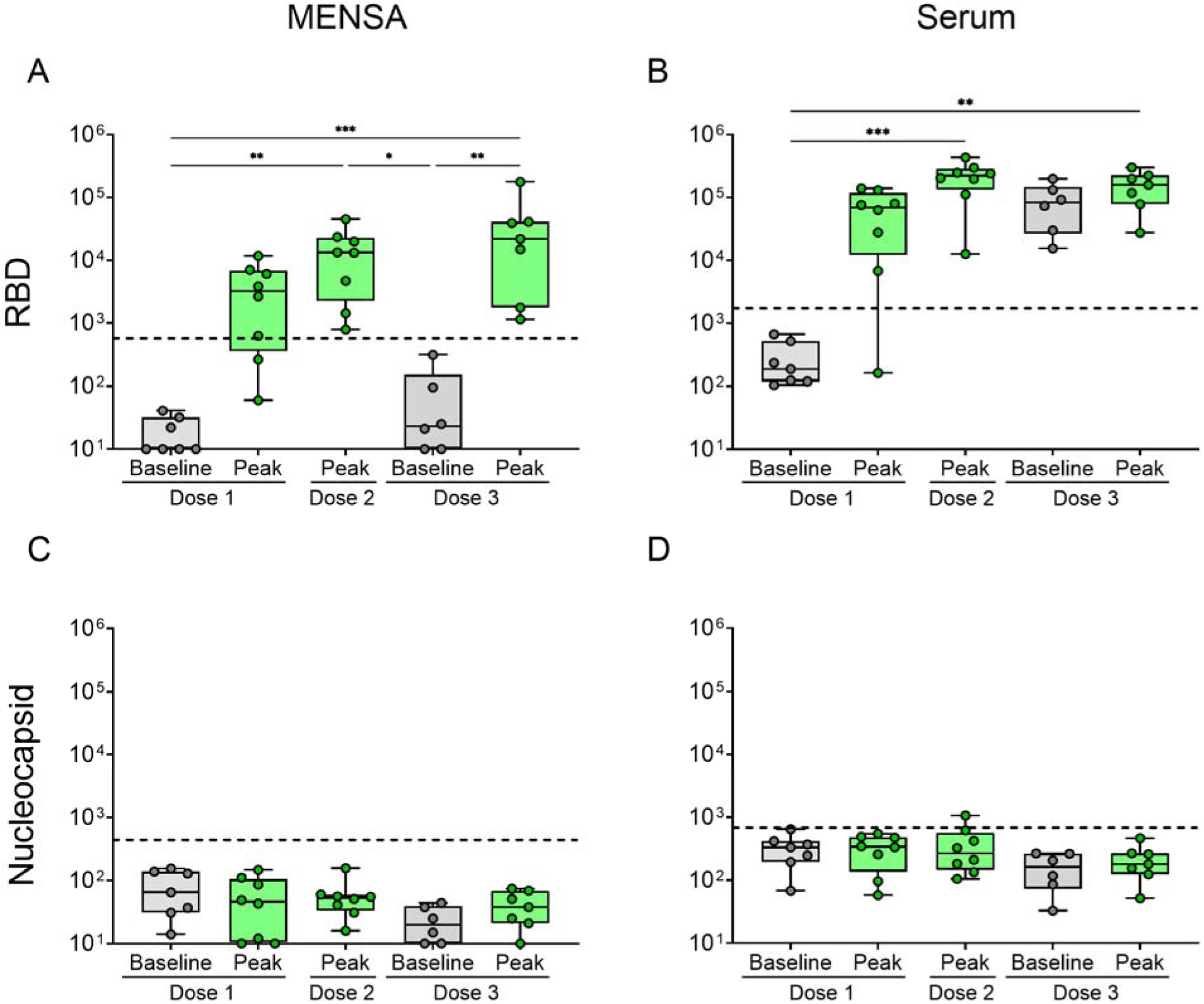
Primary vaccination responses in MENSA and serum. Dot plots show IgG antibody reactivity against S1-RBD in the MENSA (A) and serum (B) of subjects receiving their primary COVID-19 mRNA vaccination, with no prior infection. Similar results are also shown for anti-nucleocapsid IgG in the MENSA (C) and serum (D). Samples were collected from 11 healthy adults prior to vaccination (Baseline; n=7), after Dose 1 Peak (9-20 DPV; n=8), Dose 2 Peak (6-12 DPV; n=8), Dose 3 Baseline (>80 DPV dose 2 through 0 DPV dose 3; n=6), and Dose 3 Peak (4-12 DPV dose 3; n=7). All values are reported as average Net MFI (Median Fluorescent Intensity – Background). Dashed lines indicate the C_0_ threshold of positivity for each sample type and antigen as determined in Figure 1. Pair-wise comparisons were performed using the Kruskal-Wallis test in GraphPad Prism (unpaired, nonparametric test; ns p > 0.05, * p ≤ 0.05, ** p ≤ 0.01, *** p ≤ 0.001, **** p ≤ 0.0001).

Similar to serum titers, MENSA antibody levels to N are also negative providing accurate responses to only the known proteins in the vaccines (**Fig. 2C, D**).

### Longitudinal time-course of MENSA and serum after SARS2 infection and vaccination

Patients were recruited during the primary SARS2 infection or prior to the primary mRNA vaccine series. Serial blood samples were collected for MENSA and serum at the first and each subsequent SARS2 exposure events (infection and vaccination) to characterize the kinetics of the immune response during repeated exposure. In **Fig. 3**, we follow a 30-year-old Caucasian male subject from his initial mild SARS2 infection in 2020, before and after three doses of the Pfizer mRNA COVID-19 vaccine in 2021, and finally during his breakthrough Omicron infection and recovery in 2022. This subject’s first draw was collected at 9 DPSO from his primary SARS2 infection in early 2020. MENSA is positive for anti-RBD and anti-N IgG during the acute infection (**Fig. 3A,C**). The serum antibody titers are also weakly positive for anti-RBD and anti-N (**Fig. 3B,D**), as expected in most primary M/M infections. By 80 DPSO, the serum levels have increased further and remained elevated for several months while the MENSA levels rapidly decrease back to baseline. Upon three vaccine doses, the MENSA anti-RBD antibody levels rise and fall as expected whereas the serum antibodies to RBD remain positive from their previous infection and demonstrate a modest increase. Again, MENSA to the N protein is negative since it is not a component of the mRNA vaccine (**Fig. 3C**). However, serum anti-N levels remain weakly positive for several months to years after initial infection and before declining (**Fig. 3D**). In 2022, two years after the primary SARS2 infection and 8 months since his last vaccine booster vaccine, this subject had a PCR-confirmed Omicron breakthrough infection. Once again, the MENSA for anti-RBD and anti-N increases together. As expected, serum anti-N titers rise rapidly while the serum anti-RBD antibody levels, which were already high, remain elevated (**Fig. 3**). Unlike serum, the kinetics of MENSA demonstrate a rapid rise after each SARS2 exposure, whether it is due to vaccination or infection, but then decline to baseline negative values. MENSA is also highly discriminatory for spike (RBD) only after vaccination but shows a combination of spike (RBD) and N antibody levels during acute infections. We present additional kinetics spanning several years from a 24-year-old Caucasian female through three doses of the Moderna mRNA COVID-19 vaccine and an Omicron breakthrough infection (**Supplementary figure 3**) and from a 35-year-old Asian male through four doses of the Moderna mRNA COVID-19 vaccine and an Omicron breakthrough infection (**Supplementary figure 4**). In all, during SARS2 exposures from infection or vaccination, the MENSA antibody levels rise and then fall while serum titers rise after the primary exposure and stay elevated.

**Figure 3.**
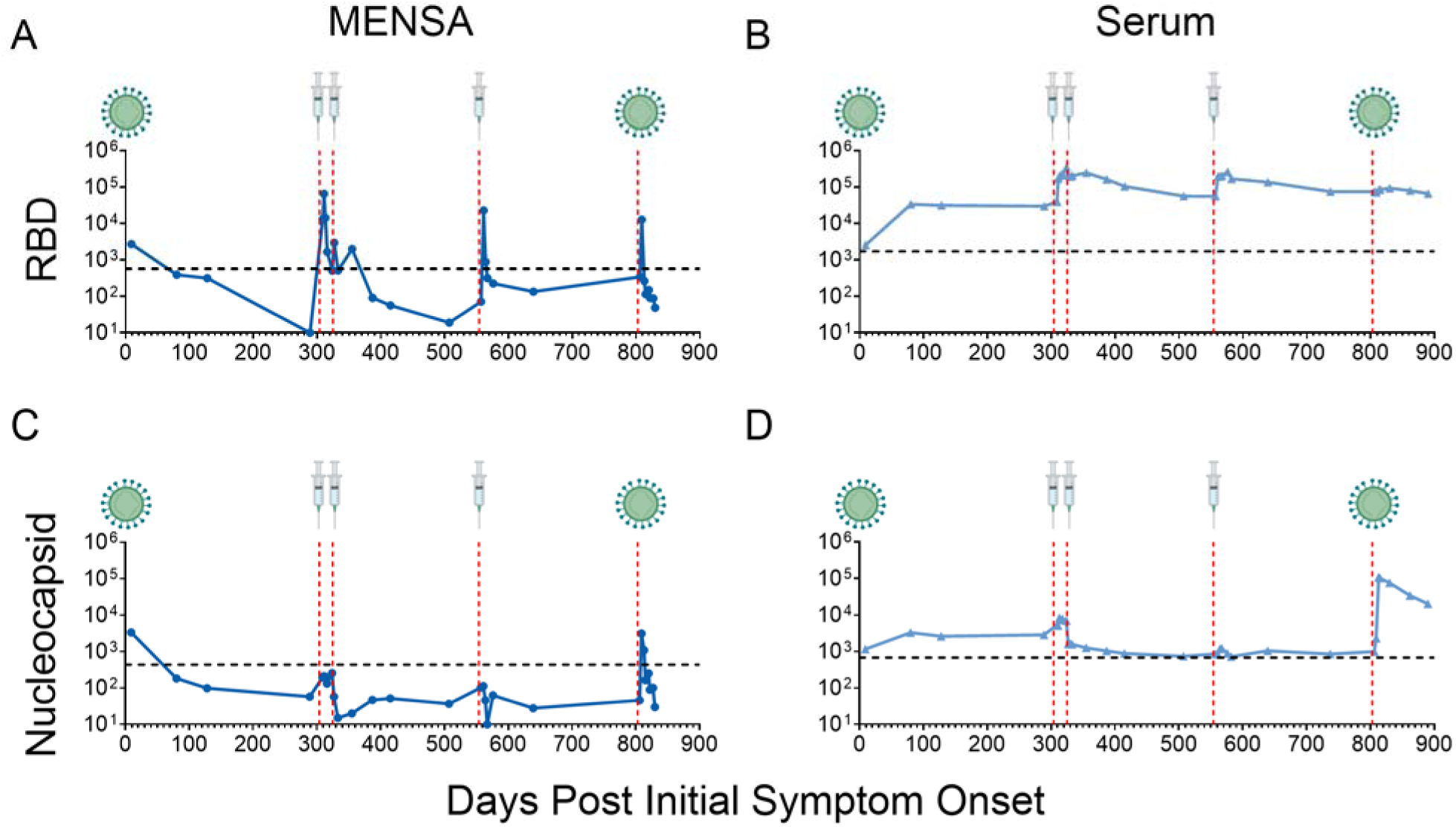
Kinetics of MENSA and serum after SARS2 infection and multiple vaccine doses. Line graphs show MENSA (dark blue) and serum (light blue) IgG antibody responses to S1-RBD (A,B) and Nucleocapsid (C,D) over time for a single patient starting with an initial primary SARS2 infection in 2020, through three doses of Pfizer COVID-19 mRNA vaccination in 2021, and a breakthrough Omicron infection in 2022. All values are reported as average Net MFI (Median Fluorescent Intensity – Background). Red vertical dashed lines represent a new exposure event. The primary and breakthrough infection events are symbolized as virions. Each vaccination dose event is symbolized as a syringe. Horizontal dashed black lines represent the C_0_ threshold of positivity for each sample and antigen combination as determined from Figure 1.

### MENSA and serum in PASC vs COVID Recovered (CR)

In addition to the 25 COVID recovered patients with no subsequent sequelae, we enrolled 61 PASC patients from the Emory Long-COVID clinic from 2021 to 2024 with self-reported symptom questionnaires at enrollment (**Fig. 4**). We reconciled symptoms collected at enrollment and with a physician chart review and follow-up. The most common self-reported symptom in this PASC cohort was enduring fatigue in 97% of patients, followed by persistent shortness of breath (SOB) in 75% with some who received new diagnoses of asthma or lung disease following their initial infection. Other common symptoms included brain fog (67%), dizziness (52%), post-exertional malaise (47%), headache (42%), chest and muscle pain (38% respectively), chronic cough (37%), depression/anxiety (37%), palpitations (32%), sleep disturbance (32%) joint pain/arthralgia (32%), and persistent loss of taste or smell (30%) (**Fig. 4).** In addition to follow-up in the Long-COVID clinic, 35% were referred for advanced neurology/neurocognitive evaluation and 58% for cardiac issues with new diagnoses of tachycardia and/or postural orthostatic tachycardia syndrome (POTS) (31%), arrhythmia/atrial fibrillation (19%), heart failure (11%), ongoing chest pain (11%), venous reflux/vein compression by doppler ultrasound (11%), dysautonomia (8%) and pericarditis/myocarditis (8%). Attempting to reconcile the 12 PASC symptom scores to better predict this long-COVID as recently reported in November 2023^6^, we calculated the average PASC score of 11.8 at enrollment for the 60 patients. Total scores equal to or greater than 12 correlated with PASC patients at 6 months, while scores less than 12 were more likely to be PASC indeterminate.

**Figure 4.**
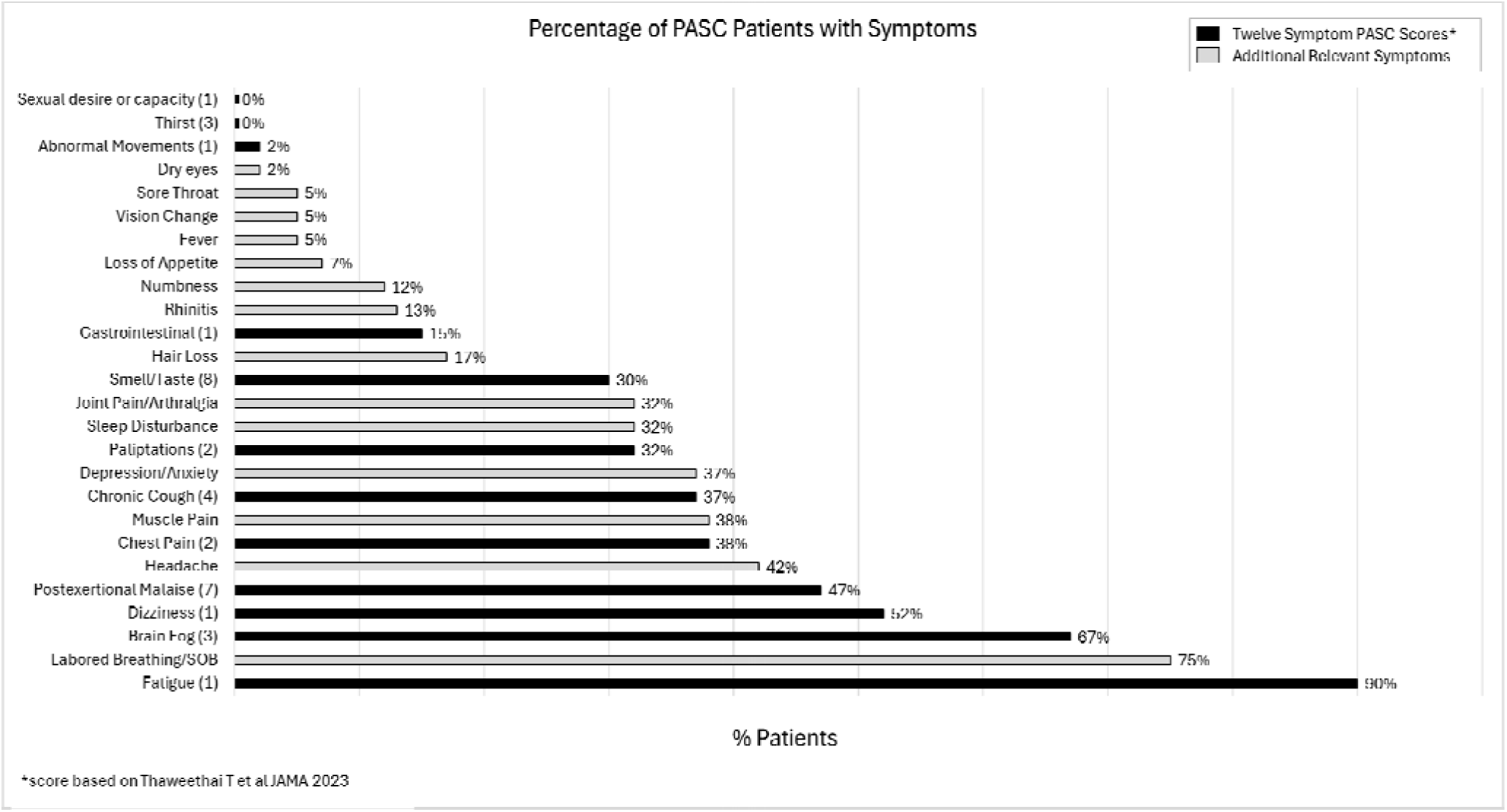
Percentage of PASC patients with symptoms. Sixty PASC patients recruited in the Emory Long-COVID clinic with self-reported symptom questionnaires at enrollment. Percentages of each symptom is shown to the right of each bar. The twelve-symptom PASC scores based on Thaweethai et al JAMA 2023 are shown in black with point scores in parentheses after each symptom.

For the initial PASC experiment, we tested for only SARS2 antigens in MENSA and serum samples prepared from the subset of 19 CR subjects mentioned above and 39 PASC patients recruited during the first year of the Long-COVID clinic at Emory University December 2020-May 2021. For the CR group, fifteen of the samples were taken directly from the Figure 1 convalescent data while four of the patients donated additional follow-up samples. All patients were enrolled from day 60-279 DPSO after their initial acute infection and prior to any COVID-19 vaccination. Of the 39 PASC patients, 56% had initial M/M acute infections and 44% had initial S/C acute infections. Of the 19 CR patients, 95% had M/M acute infections and 5% had S/C acute infections. In the 39 PASC patients, 33% still had positive MENSA for spike RBD after 60 DPSO compared to none in the CR group (**Fig. 5A**). Only 10% of PASC patients and none of the CR patients had a positive MENSA for N (**Fig. 5C**). In serum, nearly all PASC (97%) and CR (95%) patients had positive antibodies for RBD and N (**Fig. 5B,D**). One patient from each group was negative for both antigens in their serum and MENSA.

**Figure 5.**
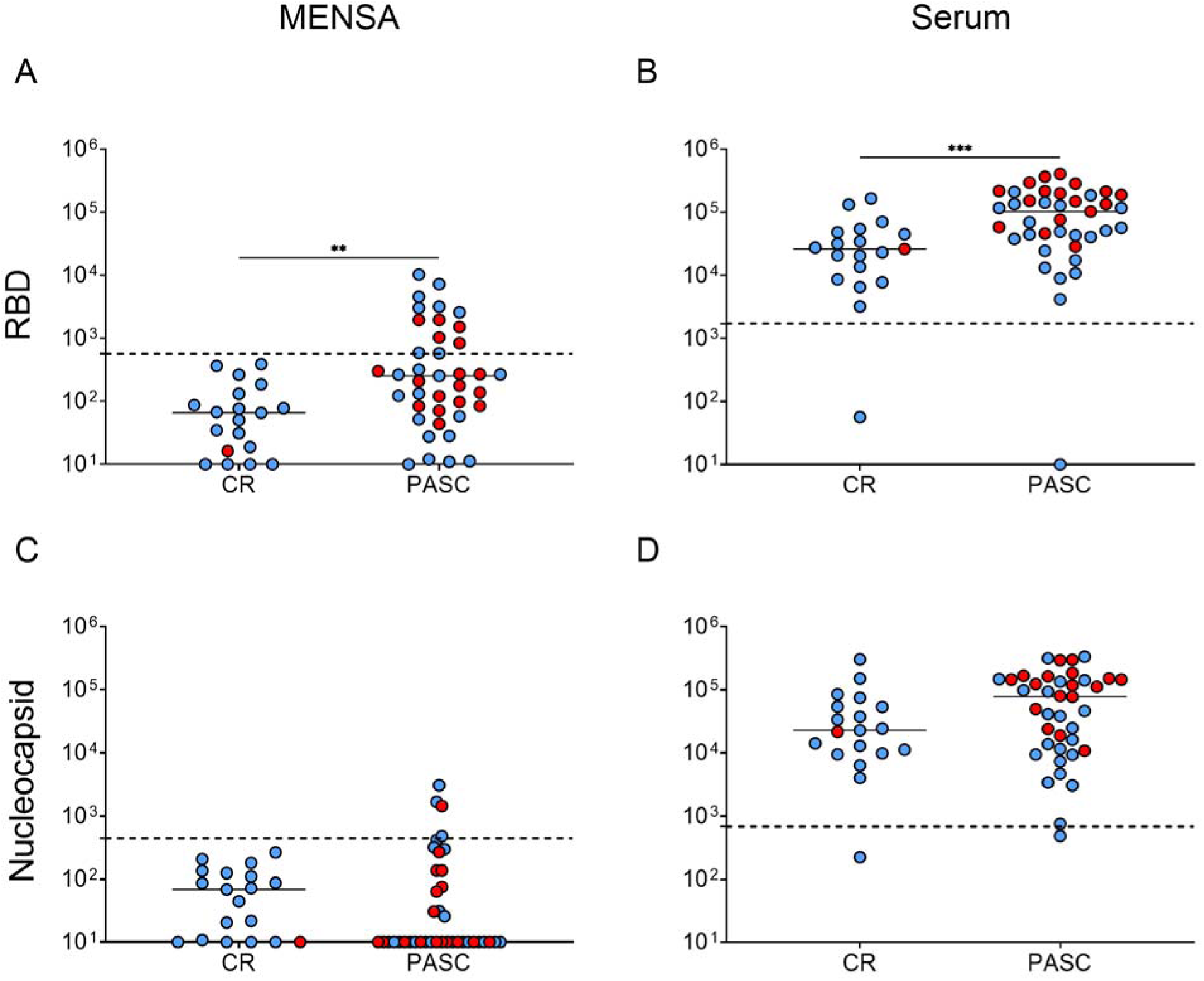
Prolonged, elevated MENSA IgG responses for SARS2 in a subset of PASC patients. Dot plots show MENSA and serum IgG antibody responses to S1-RBD (A,B) and Nucleocapsid (C,D) in samples collected between 60-279 DPSO since initial COVID-19 Wild Type infection from patients who completely recovered from their acute illness (CR; n=19) and patients who suffer PASC (n=39). Blue dots represent a Mild/Moderate acute disease severity. Red dots represent a Severe/Critical acute disease severity. All values are reported as average Net MFI (Median Fluorescent Intensity – Background). Dashed lines indicate the C_0_ threshold of positivity for each sample type and antigen combination as determined from Figure 1. Pair-wise comparisons were performed using the Mann-Whitney test in GraphPad Prism (unpaired, nonparametric test; ns p > 0.05, * p ≤ 0.05, ** p ≤ 0.01, *** p ≤ 0.001, **** p ≤ 0.0001).

### Human viral scan in MENSA of PASC and CR

For discovery of other human virus reactivation in the MENSA samples, we compared MENSAs collected from 10 PASC patients in 2021 and three CR patients using the human PhIP-seq single-end DNA sequences that were aligned to a library of reference DNA sequences of 149,259 peptides tiling protein-coding sequences from all viruses with human hosts^31,32^. Since it was early in the pandemic, the PhIP-seq was not optimized for SARS2. After quality control, we identified 227 peptides which were positive in MENSA in greater than three PASC patients and identified three additional major viruses, EBV, CMV, and HSV2 (**Fig. 6**). Thus, a new PASC MENSA assay was developed using viral antigens from SARS2, EBV, CMV, and HSV2 (see methods).

**Figure 6.**
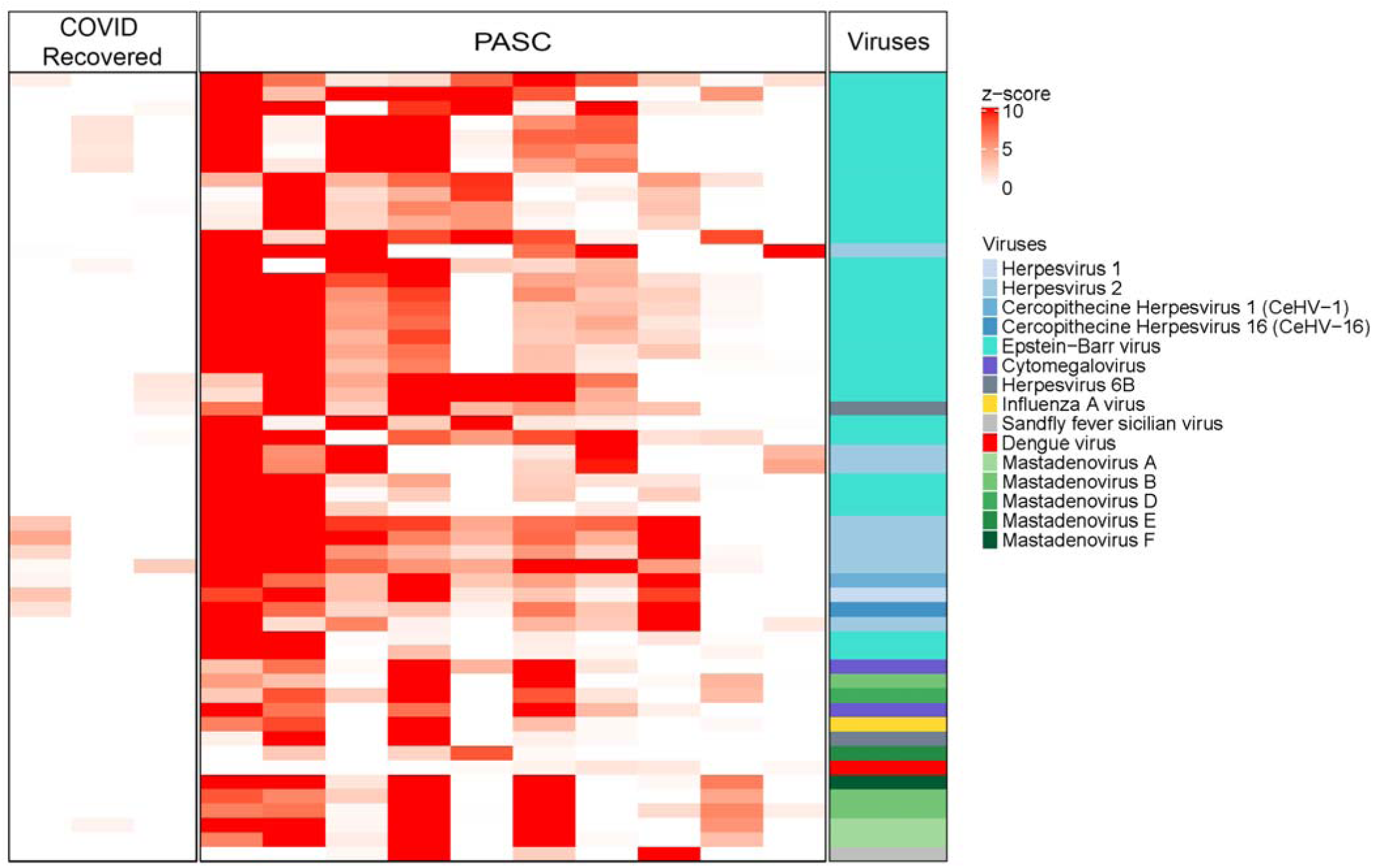
Phage immunoprecipitation sequencing (PhIP-Seq) analysis of MENSA of PASC and CR patients for discovery. PhIP-seq analysis determines the level of binding of antibodies to 149,259 peptides tiling all protein-coding sequences from viruses with human hosts in MENSA samples from three CR (left column) and 10 PASC patients (middle column). Data are presented as z-scores of the anti-viral antibodies detected. Each row represents a linear peptide of viruses that were differentially bound in PASC and CR groups. Color code identifies virus species (right column).

### MENSA for SARS2, EBV, CMV, and HSV2 in PASC and CR

In a cohort of 60 PASC patients (39 patients in 2021 and 21 patients from 2022-2024), 21 CR patients (2020-2022), and 16 healthy adult controls (2020), we tested MENSA and serum for a combined SARS2, EBV, CMV, and HSV2 IgG immunoassay. We had 23 samples from 21 CR patients since two individuals suffered repeat SARS2 infections in 2020 and 2022. As expected, all healthy controls drawn prior to SARS2 exposure were negative for SARS2 IgG in both MENSA and serum **(Fig. 7).**

**Figure 7.**
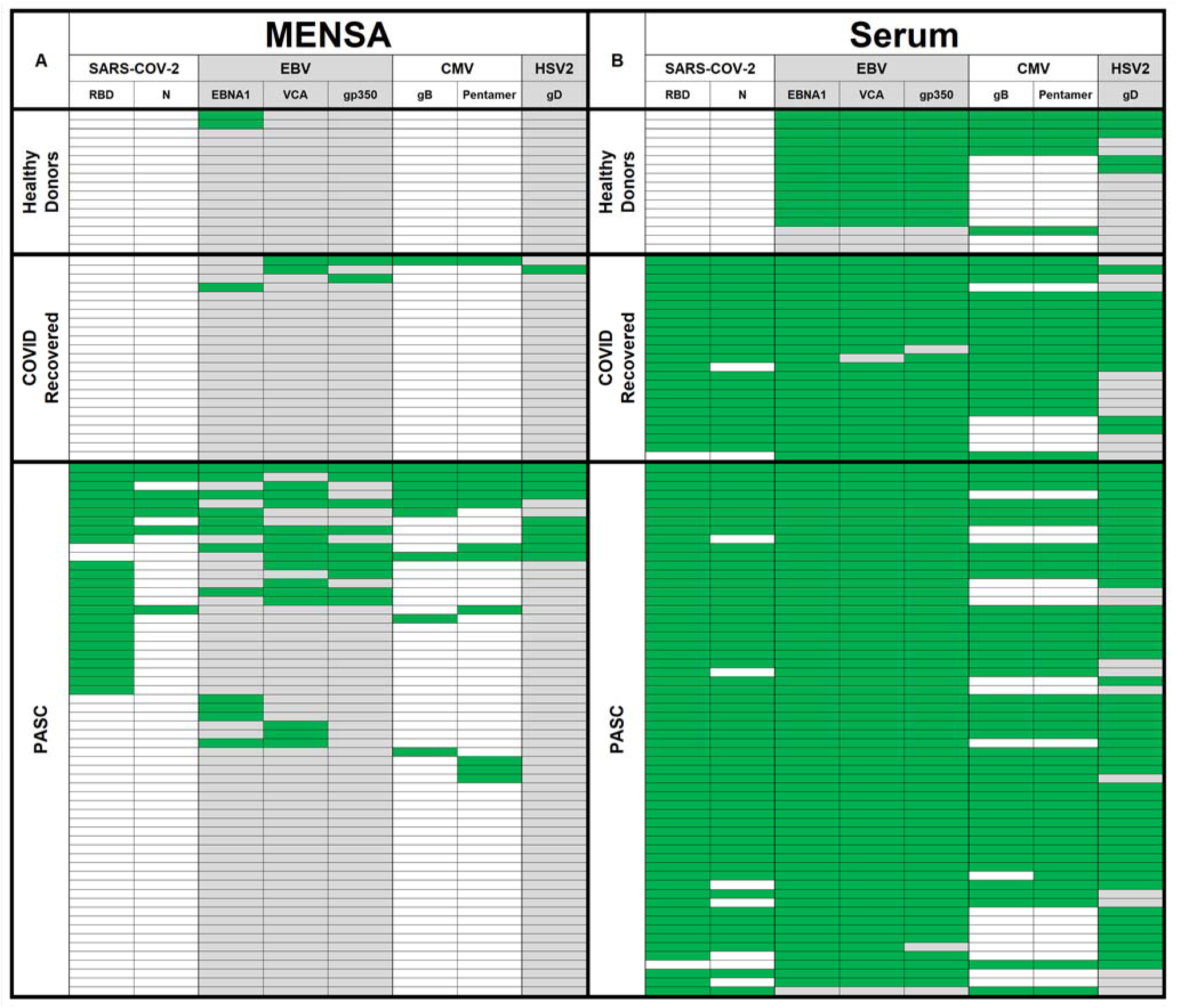
MENSA and serum in PASC, CR, and healthy adults for SARS2, EBV, CMV, and HSV2. MENSA (A) and serum (B) samples were collected from healthy donors prior to SARS2 exposure (top), COVID recovered (middle), and PASC (bottom) patients, tested for IgG reactivity against SARS2, EBV, CMV, and HSV2 antigens, and presented in a heat map. Green cells represent Net MFI values ≥ the C_0_ thresholds calculated for each sample type and antigen combination, while white or grey cells represent values below the C_0_. In MENSA, a C_0_ was calculated as the average Net MFI of 22/23 CR samples plus three standard deviations for each antigen. Each SARS2 Serum C_0_ was calculated as the average plus three standard deviations of the 16 Healthy Donor samples collected prior to SARS2 exposure. For each of the remaining viruses, three clinically confirmed negative serum samples were obtained as virus specific negative controls and the average Net MFI plus three standard deviations were used to calculate C_0_ for each antigen.

Only 2/16 (13%) were positive in the MENSA for any of the viruses tested (EBV) in the healthy control group, whereas 14/16 (88%) were positive for EBV, CMV, and/or HSV2 in the serum. In the PASC vs CR groups, we show positive MENSA for SARS2 in 24/60 (40%) of PASC patients and none in the CR (**Fig. 7A**). Nearly all PASC and CR patients are positive for the antibodies to SARS2 in the serum (98% and 96%, respectively) (**Fig. 7B**). Interestingly, the lack of correlation between MENSA and serum suggests they function as independent variables, although frequencies of plasmablasts may be linked with rise in serum titers.

When examining the latent herpes viruses, MENSA reactivity was greater overall in the PASC group than in the CR group (**Fig. 7A**). For EBV, more PASC patients had positive MENSA samples 22/60 (37%) compared to CR subjects 4/23 (17%). A MENSA test was scored positive if any one of the 3 antigens (EBNA1, VCA, and gB350) was positive. Nearly all individuals in the general population have been exposed to EBV and our results were consistent with this finding. Here, EBV serologies are positive for 59/60 (98%) in the PASC group and all 23 CR samples.

MENSAs for CMV are positive in 14/60 (23%) of the PASC patients compared to 1/23 (4%) in the CR group. Again, a test was scored positive if one of the two CMV antigens (gB or pentamer) was positive. Similar frequencies of positive CMV serology are notable in the PASC patients 43/60 (72%) versus CR individuals 18/23 (78%). For HSV2, MENSA samples are positive in 9/60 (15%) of the PASC patients, and 1/23 (4%) in the CR samples. The serum was positive in 49/60 (82%) and 12/23 (52%) of the PASC and CR samples, respectively. Overall, MENSA assays are positive in 47% (28/60) of the PASC patients for EBV, CMV, and/or HSV2 whereas only 17% (4/23) are positive in the CR cohort. In all, we identify a positive MENSA in 36/60 (60%) PASC for any of the 4 viruses (SARS2, EBV, CMV, or HSV2) compared to 4/23 (17%) for the CR group. In conclusion, a positive MENSA for SARS2 in PASC patients demonstrates ongoing new immune responses consistent with a reservoir for the persistence of SARS2 virus. Moreover, a positive MENSA for any of the 3 herpes viruses also demonstrates reactivation of latent EBV, CMV, and HSV2 identifying underlying viral triggers in this condition.

## DISCUSSION

Understanding the main viral drivers of the inflammatory and metabolic changes in patients with long-COVID has been challenging. Multiomic studies provide a wealth of information but have not identified the underlying triggers of this chronic condition. Although suggestions of viral persistence have been raised, it has been difficult to demonstrate ongoing reservoirs or reactivation of the latent virus by PCR due to the limited sensitivity of the current tests. Thus, detecting the pathogen has been challenging in patients with normal or even heightened immune responses. In this paper, we offer MENSA as a novel approach to identify the main viral drivers of long-COVID. MENSA ascertains unique immune signatures by capturing the antibodies from the circulating plasmablasts. These antibodies in the MENSA provide an immune snapshot that reveals the underlying drivers of the current illness. As proof of concept, we show that with known exposure to SARS2 by infection or vaccination, MENSA from the blood is positive. Applying these same principles, in a cohort of 60 PASC patients of whom we did not know the underlying cause, we show that 60% have a positive MENSA response against SARS2, EBV, CMV, and/or HSV2, thereby demonstrating ongoing reservoirs of SARS2 and/or reactivation of latent herpes viruses.

With first time infections, naive B cells are activated and undergo massive expansions through extrafollicular and germinal center reactions in the lymph nodes to form memory B cells and newly-minted ASC that produce antibodies. Interestingly, the majority of these ASC die, but a few successfully migrate to the bone marrow or tissue sites where they can undergo further maturation to become long-lived plasma cells (LLPC). Nearly all ASC circulating in blood are newly generated and display markers of recent proliferation such as Ki67^33,34,35^, unlike LLPC which stop proliferating. Memory B cells persist over a lifetime and differentiate into plasmablasts when re-encountering the same antigens^35^. During breakthrough or repeat infections, newly-minted ASC mostly originate from memory B cells and circulate transiently in the blood^35^. Since MENSA measures antibodies only from these newly-minted ASC and not from old LLPC, MENSA antibodies provide a unique antibody signature to reveal the cause of the present-day illness. We show that, during convalescence, the MENSA responses become negative because memory B cells are no longer differentiating into ASC and released into the blood. Thus, MENSA offers an immune snapshot to uncover the sources of the patient’s ailment.

Despite the high sensitivity of PCR testing in the nasopharyngeal swabs (NPS), blood PCR tests have limited utility for SARS2 and latent herpes viruses, such as EBV, CMV, and HSV2 due to strong T cell responses that mediate rapid viral clearance. SARS2 antigen assays have been shown to identify the spike protein, but quantities are extremely low and require ultrasensitive assays which carry a high risk of false positives^14^. Autopsies up to 230 days after acute SARS2 infection detected SARS2 RNA in multiple tissues such as the gut, central nervous system (CNS), muscle, myocardium, and the respiratory tract^36^ demonstrating viral reservoirs. Thus, measuring the MENSA has advantages over pathogen detection by PCR amplification or protein since MENSA is in the blood, agnostic to viral reservoir locations, and would not require invasive tissue sampling. Since MENSA culminates from the total newly-minted ASC traveling in the blood during acute illness, knowledge of the viral reservoir location is not necessary since the MENSA reveals infections in deep-seated sites similar to a liquid biopsy.

Breakthrough or repeat infections can be diagnosed with serum assays, but they typically require serial blood samples during acute infection and convalescence for comparison. Another advantage of the MENSA over serum is that only a single blood sample during illness is needed. The decline of MENSA to negative levels after infection or vaccination demonstrates its clinical utility in measuring secondary or breakthrough infections.

Specificity of MENSA antibodies are also exact in that they can distinguish infection from vaccination based on spike and the nucleocapsid proteins in some patients. This specificity and sensitivity along with the kinetics make MENSA an ideal diagnostic platform to reveal viral triggers that were previously difficult to measure with just serum or PCR tests. The MENSA diagnostic would be the first of its kind to understand the main viral drivers of this chronic disease.

MENSA antibodies are expected to peak within days after exposure, and then quickly decline back to baseline within a month after the infection has resolved. Interestingly, CR MENSA does not revert to pre-pandemic baseline levels and these mechanisms are not clear. Perhaps non-specific plasma antibody binding to monocytes in the MENSA cultures may be the reason and will require more studies. Another possibility is low-level bystander responses which have been suggested^35^ to explain the difference between pre- and post-pandemic samples. Interestingly, even when using the post-pandemic MENSA samples as controls, 60% of the PASC patients have higher SARS2, EBV, CMV, and HSV2 responses in the MENSA.

Autoantigen triggers have also been implicated in PASC patients, and so we tested MENSA from a limited number of PASC patients for autoantigens using the PhIP-seq human peptidome library which consists of 605,656 peptides tiling protein-coding sequences, splice variants, non-coding open reading frames, and endogenous retroviral sequences in the human genome ^37,38^. No differences were observed between the PASC and CR MENSA against the human peptidome (our unpublished results). However, a larger number of patients using the 3-D conformational epitopes of the human proteome may be needed to definitively rule out MENSA responses to autoantigens. Since the original PhIP-seq assays used linear viral peptides, we may also consider a panel of 3-D conformational epitopes to identify important unique immune signatures for viruses that infect humans to ensure comprehensive testing for other viruses.

There are several limitations of this study. One is that we do not have longitudinal samples from the PASC patients and thus, it is unclear how consistent the MENSA responses are in the patients with chronic illness over time. Second, large clinical trials are needed to evaluate responses to anti-viral therapies in MENSA positive patients identified with SARS2 persistence and reactivation of EBV, CMV, or HSV2 infections. Finally, the utility of MENSA may be challenging in immunocompromised patients since the MENSA requires B cell activation to form new ASC.

The real-time immune snapshots provided by MENSA may be leveraged to inform therapeutic strategies and successful treatment of chronically ill PASC patients. Whether MENSA can also be useful to identify persistence of viral reservoirs in other chronic illnesses such as multiple sclerosis, HIV, myalgic encephalomyelitis/chronic fatigue syndrome (ME/CFS) and other infections with post-sequelae are yet to be determined. For example, EBV was recently implicated in multiple sclerosis^39^. Interestingly, infection with SARS2 is associated with increased susceptibility and severity of neurodegenerative disorders, such as Alzheimer’s disease, Parkinson’s disease, and dementia but interpreting these correlations has been difficult ^40–42^. If MENSA can lead to early diagnosis of these chronic neurodegenerative disorders or help identify the cause of these disease flares, perhaps treatments may prove more effective in preventing progression and severity of these pathological conditions.

PASC can include symptoms, such as dyspnea, fatigue, and depression^43^, and serious clinical indications, such as cardiovascular disease or diabetes^44,45^. Recent studies also suggest increased risk for autoimmune inflammatory rheumatic diseases in PASC and CR patients^46–48^. Future clinical trials are necessary to perform proof-of-concept studies where MENSA data could be used to inform treatment modalities for mitigating symptoms associated with SARS2 viral persistence, reactivation of viruses, reactivation of viruses in other chronic illnesses, or early activation of other chronic illnesses.

In summary, MENSA is a novel immune diagnostic which captures unique signatures of the early-minted ASC in the blood to reveal the cause of illness. In chronic conditions such as PASC where serum antibody titers are high, MENSA is an independent matrix that identifies persistence of SARS2 viruses or antigens and can also recognize the reactivation of latent herpes viruses, such as EBV, CMV, and HSV2 in 60% of patients. This host immune snapshot reveals the fundamental drivers of viral persistence and reactivation in this chronic disease.

## METHODS online

### Subject Enrollment and Sample Collection

#### Patient enrollment

We enrolled 241 adults between 2020 and 2024 at Emory University in Atlanta, GA and collected blood to generate the MENSA matrix and serum samples (Table 1). In 2020, we enrolled 110 adults with PCR-positive NPS during their primary SAR2 infection. Fifty-four adults were outpatients with mild/moderate (M/M) disease as defined by the NIH criteria^28^, and 56 adults had severe/critical (S/C) illness in the Intensive Care Units. We also enrolled 60 healthy adults with no known exposure to SARS2 early during the pandemic with blood samples collected between March and June 2020 during the initial lockdown as the healthy adult controls without prior SARS2 exposure. Eleven vaccinated subjects who had no prior SARS2 infection were enrolled before and during their primary two-dose mRNA vaccine series (Pfizer n=5, Moderna n=6) and drawn again before and after their third booster dose. We also enrolled 3 adults experiencing Omicron breakthrough infections between December 2021 and June 2022.

Finally, we enrolled 61 long-COVID or PASC patients from the Emory Long-COVID Clinic which was started in January 2021 until February 2024 (Fig. 4). Initially, 40 patients were enrolled in 2021 (Figs. 5, 6, 7) and an additional 21 patients were enrolled in 2022-2024 (Fig. 7). All patients filled patient-reported symptom questionnaires. Samples from nine of the initial 39 PASC patients from Fig. 5 and one additional PASC patient recruited in 2021 were sent to Immune ID for PhIP-seq analysis along with samples from three COVID Recovered patients (1/3 CR from initial convalescent cohort in Fig 1; 2/3 new CR patients). All blood samples were collected under the Emory University Institutional Review Board–approved protocols.

##### MENSA Preparation

Medium enriched for newly synthesized antibodies (MENSA) was generated by isolating, washing, and culturing antibody-secreting cells (ASC)-containing peripheral blood mono-nuclear cells (PBMC) from blood using a modified procedure previously described ^23^. Peripheral blood samples were collected in sodium heparin tubes and PBMC were isolated by centrifugation (1,000 ×g; 10 min) using Lymphocyte Separation Media (Corning) and Leucosep tubes (Greiner Bio-One). Five washes with RPMI-1640 (Corning) were performed to remove serum immunoglobulins (800 x g; 5 min) with erythrocyte lysis (3 mL; 3 min), and harvested PBMCs were cultured at 10^6^ cells/mL in R10 Medium (RPMI-1640, 10% Sigma FBS, 1% Gibco Antibiotic/Anti-mycotic) for 24 h at 37° C and 5% CO_2_. After incubation, the cell suspension was centrifuged (800 ×g; 5 min), and the supernatant (MENSA) was separated from the PBMC pellet, aliquoted, and stored at -80°C for testing.

##### Serum Preparation

Whole blood was collected and incubated at room temperature for at least 30 minutes. The clot was discarded, and the remaining serum supernatant was centrifuged (800xg; 10 min), aliquoted and stored at -80 °C for testing.

##### Antigen Selection and Multiplex Immunoassays

Antigens of interest were selected from literature, coupled to Luminex MagPlex Microspheres of spectrally distinct regions via carbodiimide coupling, and tested for antigen specific IgG reactivity against patient samples as previously described ^29^.

#### SARS2 antigens

SARS-CoV-2 Spike S1 Receptor Binding Domain (RBD; catalog no. Z03483; expressed in HEK293 cells) and Nucleocapsid protein (N; catalog no. Z03480; expressed in Escherichia coli), were purchased from GenScript. S1 (catalog no. S1N-C52H3; HEK293), S2 (catalog no. S2N-C52H5; HEK293) and S1 N-terminal domain (NTD; catalog no. S1D-C52H6; HEK293) were purchased from ACROBiosystems. The C-terminus sequence of ORF3a (Accession: QHD43417.1, amino acids 134-275 plus N-terminal His6-Tag) was sent to Genscript for custom protein expression in E. coli. Each protein was expressed with an N-terminal His6-Tag to facilitate purification, at least 90% pure, and appeared as a predominant single band on SDS-PAGE analysis.

#### EBV, CMV, and HSV2 antigens

EBV, CMV, and HSV2 antigens were carefully selected for antigenicity based on previous reports^49–54^. The following proteins were used: EBV EBNA1 protein from Abcam (produced in E. coli, N-Terminus His Tag, CAT#ab138345); EBV VCA p18 from RayBiotech (produced in E. coli, CAT#227-20127); EBV gp350 protein from AcroBiosystems (produced in HEK293 cells, His Tag, MALS verified, CAT#GP0-E52H6); CMV glycoprotein B from AcroBiosystems (strain AD169, expressed from HEK293 cells, His Tag, MALS verified, CAT#CMB-V52H4); CMV gH pentamer complex, consisting of gH, gL, UL128, UL130 and UL131A proteins, produced in mammalian HEK293 cells from The Native Antigen Company (CAT#CMV-PENT); HSV2 envelope glycoprotein D from AcroBiosystems (gD, expressed HEK293 cells, His Tag, MALS verified, CAT#GLD-V52H4).

#### Serum and MENSA assays for SARS2 and other viruses

Serum samples were tested at 1:500 dilution in assay buffer (1XPBS, 1% BSA) while MENSA samples were tested neat with no dilution. Results were analyzed on a Luminex FLEXMAP 3D instrument. Median fluorescent intensity (MFI) using phycoerythrin-conjugated detection antibodies (Goat Anti-Human IgG-PE, Southern Biotech cat. #2040-09) was measured for each sample using the Luminex xPONENT software on Enhanced PMT setting. The background value of assay buffer or R10 media was subtracted from the serum or MENSA results, respectively, to obtain MFI minus background (Net MFI). All samples were tested in duplicate and the average of the two results were used for analysis. In the initial SARS2 assay, all SARS2 protein bound microparticles were run together as a six-bead SARS2 solution. Serum C_0_ values of positivity were calculated as the average plus five standard deviations of the Healthy Control population (N=60) for each antigen (RBD: 1724; N: 682). MENSA C_0_ positivity values were calculated as the average plus three standard deviations of the Contemporary Controls (COVID Recovered N=19 (CR) group described above) for each antigen (RBD: 570; N: 441).

Later, the assay was modified to contain only RBD, N, S1, and S2 from SARS2 (NTD and ORF3a dropped) and also included the addition of all EBV, CMV, and HSV2 antigens, for a combined multi viral 10-antigen bead assay. All new C_0_s were calculated based on the new assay data in Figure 7. MENSA C_0_s were calculated as the average plus three standard deviations of 22/23 CR samples (RBD: 404; N: 266; EBNA1: 475; VCA: 426; gp350: 523; gB: 298; Pentamer: 455; gD: 308). One sample was excluded from the MENSA C_0_ calculation due to multiple antigen reactivity measuring greater than 10 times the median value of the entire group. SARS2 Serum C_0_ was calculated as the average plus three standard deviations of the 16 Healthy Donor samples collected prior to SARS2 exposure. Since the majority of the population is expected to be positive for EBV, CMV, and/or HSV2 antibodies in their serum, we obtained de-identified clinically confirmed negative sera from the Emory clinical laboratory. For each virus, three confirmed negative serum samples were used. The average Net MFI plus three standard deviations was calculated for each antigen and used as the C_0_ threshold for positivity (RBD: 410; N: 564; EBNA1: 3,578; VCA: 593; gp350: 228; gB: 778; Pentamer: 769; gD: 858).

#### Phage immunoprecipitation sequencing and analysis

We constructed a custom T7 bacteriophage library consisting of 149,259 peptides tiling all protein-coding sequences from viruses with human hosts^31,32^. Viral sequences were downloaded from Uniprot, collapsed on 90% identity, and bioinformatically parsed into 90 amino-acid peptide tiles with 45 amino-acid overlaps between adjacent tiles. Healthy and Covid patients’ plasma or serum and matched MENSA reactivities were profiled using phage immunoprecipitation and sequencing (PhIP-seq). MENSA samples were profiled in duplicate and plasma/serum samples in triplicate. PhIP-seq was performed as previously described with some modifications^32^. T7 bacteriophage libraries were aliquoted into 96-well plates and incubated with 20μl each of protein A and G Dynabeads on a rotator for 4 h at room temperature. Next, plates were placed on a magnet and supernatants were transferred to a fresh 96-well plate, to which we added patient plasma containing 2μg of total IgG, and continued with the immunoprecipitation and washing steps, as previously described. Following the washes, protein A and protein G Dynabeads were resuspended in PCR master mix, amplified with 16 rounds of PCR, SPRI cleaned to remove primers, and indexed for sequencing with 8 rounds of PCR with primers containing Illumina p5 and p7 barcodes. NGS libraries were quantified on a Tapestation4200 and normalized for sequencing on Illumina Nextseq 2000 or Novaseq 6000 instruments. Each sequencing library received a minimum of 3M reads.

PhIP-seq single-end DNA sequences were aligned to a library of reference DNA sequences (149,259 75bp for viral) with the bowtie2 aligner (v2.0) using end-to-end matching. Read counts were summarized using samtools (v1.14) and collated into a counts matrix. The raw counts were converted to counts per million (CPM) using the ‘cpm‘ function from the R package edgeR (v3.36.0). CPM values for healthy controls were summarized by computing the peptide-wise mean and standard deviation across all healthy control samples. CPM values for each patient sample were collapsed by computing the peptide-wise minimum across technical replicates.

Peptide-wise z-scores were then computed as:

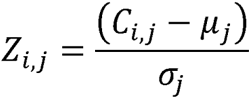

where z_i,j_ is the z-score for patient *i*, peptide *j*; C_i,j_ is the minimum CPM for patient *i*, peptide *j; μ_j_* is the mean of peptide *j* in the healthy control samples, and *σ_j_* is the standard deviation of peptide *j* in the healthy control samples. For each patient, hits were identified as those peptides with *c_i,j_* ≥ 10 AND *z_i,j_* ≥ 10.

## Data Analysis

All graphs were designed using GraphPad Prism, Excel, R studio, BioRender, and/or Adobe Photoshop. Comparisons among and between groups in figures 1,2,5 and supplemental figures 1 and 2 were calculated by Kruskal-Wallis tests, Mann-Whitney tests, and/or receiver-operating characteristic (ROC) curves using GraphPad Prism. To avoid overrepresentation of negative values on log scales, all values less than 10 Net MFI were replaced with 10 for figures 1-3 and 5. All calculations and statistics were performed using real values.

## Author Contributions

FEL, JLD, and IS designed and supervised the study. NSH, FEL, JLD, SMY, AMP, and IS wrote and/or edited the manuscript. NSH, FEL, and JLD developed and optimized the multiplex immunoassays and MENSA technology used in this study. NSH, AMP, HQ, FA, VC, AC, and MCW performed experiments and analyzed the data. AMP, HQ, FA, AC, JP, DCN, IH, CYK, SK, BS, EW, HG, DS, KSC, MCM, and NSH collected and processed the samples. TN and VB performed the PhIP-seq experiments. VC, RTA, PAL, MRH, MCR, ADT, AWD, RG, RP, KV, SU, JV, AJ, SL, SNL, AND, JBO, and RPR recruited and managed patient enrollment. DNA, JDR, and MCH provided serum samples. All authors contributed to the manuscript revision, read, and approved the submitted version.

## Data Availability

All data produced in the present study will be available upon reasonable request to the authors.

## Acknowledgments

This work was supported by National Institute of Allergy and Infectious Diseases, National Institutes of Health 3P01AI125180-05S1, R01AI121252, R01AI 172254, P01A1078907, U01AI045969, U19AI109962, U54CA260563, T32HL116271-07, and NIGMS 2T32GM095442, NIH Department of Health and Human Services/Public Health Services: 5T32AI74492-14.

## REFERENCES

1. Omer, S.B., Malani, P. & Del Rio, C. The COVID-19 Pandemic in the US: A Clinical Update. JAMA 323, 1767–1768 (2020).

2. Organization, W.H. COVID-19 weekly epidemiological update, 29 December 2020. (2020).

3. El Sahly, H.M., et al. Efficacy of the mRNA-1273 SARS-CoV-2 Vaccine at Completion of Blinded Phase. N Engl J Med 385, 1774–1785 (2021).

4. Walsh, E.E., et al. Safety and Immunogenicity of Two RNA-Based Covid-19 Vaccine Candidates. The New England journal of medicine 383, 2439–2450 (2020).

5. Kumar, S., Thambiraja, T.S., Karuppanan, K. & Subramaniam, G. Omicron and Delta variant of SARS-CoV-2: A comparative computational study of spike protein. J Med Virol 94, 1641–1649 (2022).

6. Thaweethai, T., et al. Development of a Definition of Postacute Sequelae of SARS-CoV-2 Infection. JAMA 329, 1934–1946 (2023).

7. Post COVID-19 condition (Long COVID).

8. Woodruff, M.C., et al. Chronic inflammation, neutrophil activity, and autoreactivity splits long COVID. Nat Commun 14, 4201 (2023).

9. Klein, J., et al. Distinguishing features of long COVID identified through immune profiling. Nature 623, 139–148 (2023).

10. Su, Y., et al. Multiple early factors anticipate post-acute COVID-19 sequelae. Cell 185, 881–895 e820 (2022).

11. Wong, A.C., et al. Serotonin reduction in post-acute sequelae of viral infection. Cell 186, 4851–4867 e4820 (2023).

12. Liew, F., et al. Large-scale phenotyping of patients with long COVID post-hospitalization reveals mechanistic subtypes of disease. Nat Immunol 25, 607–621 (2024).

13. Cervia-Hasler, C., et al. Persistent complement dysregulation with signs of thromboinflammation in active Long Covid. Science 383, eadg7942 (2024).

14. Swank, Z., et al. Persistent Circulating Severe Acute Respiratory Syndrome Coronavirus 2 Spike Is Associated With Post-acute Coronavirus Disease 2019 Sequelae. Clin Infect Dis 76, e487–e490 (2023).

15. Peluso, M.J., et al. Chronic viral coinfections differentially affect the likelihood of developing long COVID. J Clin Invest 133(2023).

16. Carter, M.J., Mitchell, R.M., Meyer Sauteur, P.M., Kelly, D.F. & Truck, J. The Antibody-Secreting Cell Response to Infection: Kinetics and Clinical Applications. Front Immunol 8, 630 (2017).

17. Halliley, J.L., et al. Long-Lived Plasma Cells Are Contained within the CD19(-)CD38(hi)CD138(+) Subset in Human Bone Marrow. Immunity 43, 132–145 (2015).

18. Lee, F.E., et al. Circulating human antibody-secreting cells during vaccinations and respiratory viral infections are characterized by high specificity and lack of bystander effect. J Immunol 186, 5514–5521 (2011).

19. Oh, I., et al. Tracking Anti-Staphylococcus aureus Antibodies Produced In Vivo and Ex Vivo during Foot Salvage Therapy for Diabetic Foot Infections Reveals Prognostic Insights and Evidence of Diversified Humoral Immunity. Infect Immun 86(2018).

20. Lee, F.E., Falsey, A.R., Halliley, J.L., Sanz, I. & Walsh, E.E. Circulating antibody-secreting cells during acute respiratory syncytial virus infection in adults. J Infect Dis 202, 1659–1666 (2010).

21. Wrammert, J., et al. Broadly cross-reactive antibodies dominate the human B cell response against 2009 pandemic H1N1 influenza virus infection. The Journal of experimental medicine 208, 181–193 (2011).

22. Wrammert, J., et al. Rapid cloning of high-affinity human monoclonal antibodies against influenza virus. Nature 453, 667–671 (2008).

23. Haddad, N.S., et al. Novel immunoassay for diagnosis of ongoing Clostridioides difficile infections using serum and medium enriched for newly synthesized antibodies (MENSA). J Immunol Methods 492, 112932 (2021).

24. Haddad, N.S., et al. Detection of Newly Secreted Antibodies Predicts Non-recurrence in Primary Clostridioides difficile Infection. J Clin Microbiol, jcm0220121 (2022).

25. Kyu, S., et al. Diagnosis of Streptococcus pneumoniae infection using circulating antibody secreting cells. PLoS One 16, e0259644 (2021).

26. Muthukrishnan, G., et al. A Bioinformatic Approach to Utilize a Patient’s Antibody-Secreting Cells against Staphylococcus aureus to Detect Challenging Musculoskeletal Infections. Immunohorizons 4, 339–351 (2020).

27. Haddad, N.S., et al. Circulating antibody-secreting cells are a biomarker for early diagnosis in patients with Lyme disease. PloS one 18, e0293203 (2023).

28. Panel, C.-T.G. Coronavirus Disease 2019 (COVID-19) Treatment Guidelines. (National Institutes of Health, 2020).

29. Haddad, N.S., et al. One-Stop Serum Assay Identifies COVID-19 Disease Severity and Vaccination Responses. Immunohorizons 5, 322–335 (2021).

30. Woodruff, M., et al. Critically ill SARS-CoV-2 patients display lupus-like hallmarks of extrafollicular B cell activation. medRxiv (2020).

31. Nakagawa, S. & Takahashi, M.U. gEVE: a genome-based endogenous viral element database provides comprehensive viral protein-coding sequences in mammalian genomes. Database (Oxford) 2016(2016).

32. Xu, G.J., et al. Viral immunology. Comprehensive serological profiling of human populations using a synthetic human virome. Science 348, aaa0698 (2015).

33. Nguyen, D.C., et al. Majority of human circulating IgG plasmablasts stop blasting in a cell-free pro-survival culture. Scientific reports 14, 3616 (2024).

34. Magri, G., et al. Human Secretory IgM Emerges from Plasma Cells Clonally Related to Gut Memory B Cells and Targets Highly Diverse Commensals. Immunity 47, 118–134 e118 (2017).

35. Phad, G.E., et al. Clonal structure, stability and dynamics of human memory B cells and circulating plasmablasts. Nat Immunol 23, 1076–1085 (2022).

36. Stein, S.R., et al. SARS-CoV-2 infection and persistence in the human body and brain at autopsy. Nature 612, 758–763 (2022).

37. Koren, I., et al. The Eukaryotic Proteome Is Shaped by E3 Ubiquitin Ligases Targeting C-Terminal Degrons. Cell 173, 1622–1635 e1614 (2018).

38. Larman, H.B., et al. Autoantigen discovery with a synthetic human peptidome. Nat Biotechnol 29, 535–541 (2011).

39. Lanz, T.V., et al. Clonally expanded B cells in multiple sclerosis bind EBV EBNA1 and GlialCAM. Nature 603, 321–327 (2022).

40. Bedran, D., Bedran, G. & Kote, S. A Comprehensive Review of Neurodegenerative Manifestations of SARS-CoV-2. Vaccines (Basel) 12(2024).

41. 41. Grant, R.A., et al. Prolonged exposure to lung-derived cytokines is associated with activation of microglia in patients with COVID-19. JCI Insight 9(2024).

42. Rippee-Brooks, M.D., et al. Viral Infections, Are They a Trigger and Risk Factor of Alzheimer’s Disease? Pathogens 13(2024).

43. Mizrahi, B., et al. Long covid outcomes at one year after mild SARS-CoV-2 infection: nationwide cohort study. BMJ 380, e072529 (2023).

44. Xie, Y. & Al-Aly, Z. Risks and burdens of incident diabetes in long COVID: a cohort study. Lancet Diabetes Endocrinol 10, 311–321 (2022).

45. Xie, Y., Xu, E., Bowe, B. & Al-Aly, Z. Long-term cardiovascular outcomes of COVID-19. Nat Med 28, 583–590 (2022).

46. Chang, R., et al. Risk of autoimmune diseases in patients with COVID-19: A retrospective cohort study. EClinicalMedicine 56, 101783 (2023).

47. Syed, U., et al. Incidence of immune-mediated inflammatory diseases following COVID-19: a matched cohort study in UK primary care. BMC Med 21, 363 (2023).

48. Tesch, F., et al. Incident autoimmune diseases in association with SARS-CoV-2 infection: a matched cohort study. Clin Rheumatol 42, 2905–2914 (2023).

49. Middeldorp, J.M. Epstein-Barr Virus-Specific Humoral Immune Responses in Health and Disease. Current topics in microbiology and immunology 391, 289–323 (2015).

50. Karsten, C.B., et al. Evolution of functional antibodies following acute Epstein-Barr virus infection. PLoS pathogens 18, e1010738 (2022).

51. Wiegers, A.K., Sticht, H., Winkler, T.H., Britt, W.J. & Mach, M. Identification of a neutralizing epitope within antigenic domain 5 of glycoprotein B of human cytomegalovirus. Journal of virology 89, 361–372 (2015).

52. Lilleri, D., et al. Fetal human cytomegalovirus transmission correlates with delayed maternal antibodies to gH/gL/pUL128-130-131 complex during primary infection. PloS one 8, e59863 (2013).

53. Macagno, A., et al. Isolation of human monoclonal antibodies that potently neutralize human cytomegalovirus infection by targeting different epitopes on the gH/gL/UL128-131A complex. Journal of virology 84, 1005–1013 (2010).

54. Belshe, R.B., et al. Correlate of immune protection against HSV-1 genital disease in vaccinated women. The Journal of infectious diseases 209, 828–836 (2014).

