## Supplementary figures for "MENSA, a Media Enriched with Newly Synthesized Antibodies, to Identify SARS-CoV-2 Persistence and Latent Viral Reactivation in Long-COVID"

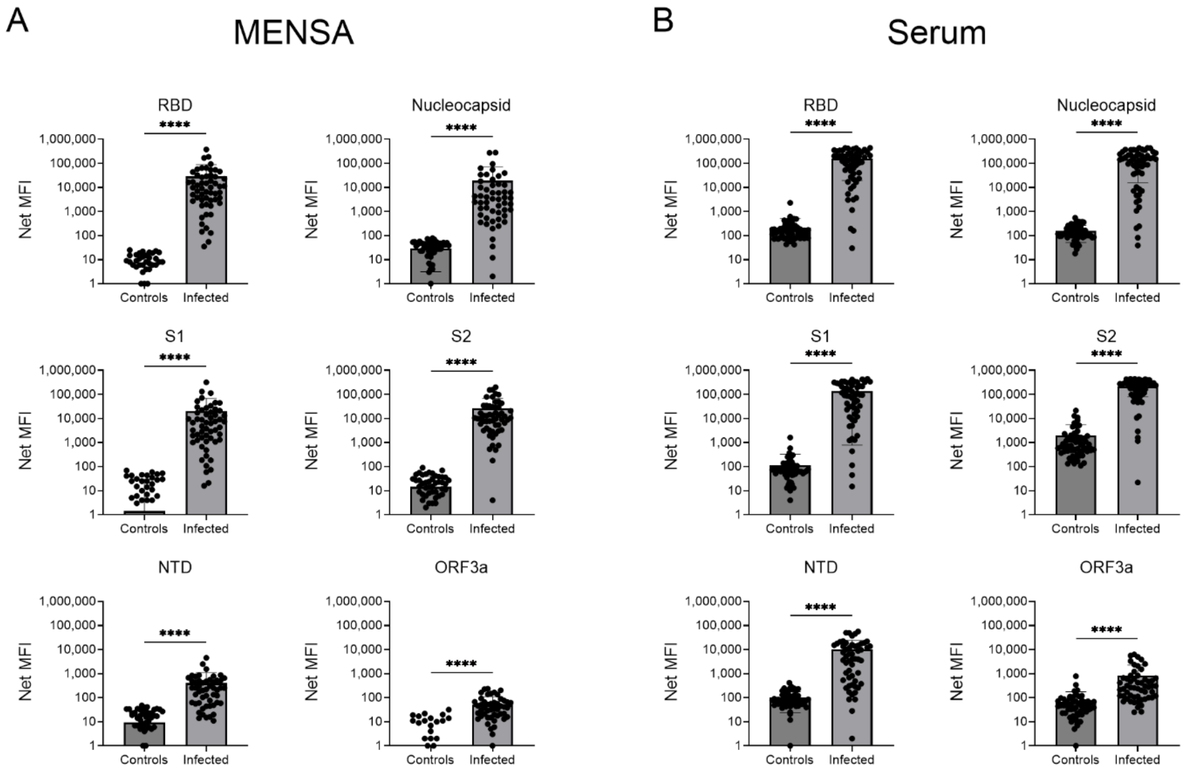


**Supplementary Figure 1. Anti-SARS-CoV-2 IgG levels are higher in the MENSA and serum of infected patients than in unexposed healthy controls.** MENSA (A) and serum (B) samples were collected from healthy adults during the early pandemic period, prior to any SARS-CoV-2 exposure, and from patients within 30 days post symptom onset (DPSO) of their primary wild type infections in 2020. All samples were tested for IgG reactivity against four spike-associated proteins (RBD, S1, S2, NTD) and two non-spike proteins (Nucleocapsid, ORF3a). All values are reported as average Net MFI (Median Fluorescent Intensity – Background). Pair-wise comparisons were performed using the Mann-Whitney test in GraphPad Prism (unpaired, nonparametric test; ns $p > 0.05$, * $p \leq 0.05$, ** $p \leq 0.01$, *** $p \leq 0.001$, **** $p \leq 0.0001$).

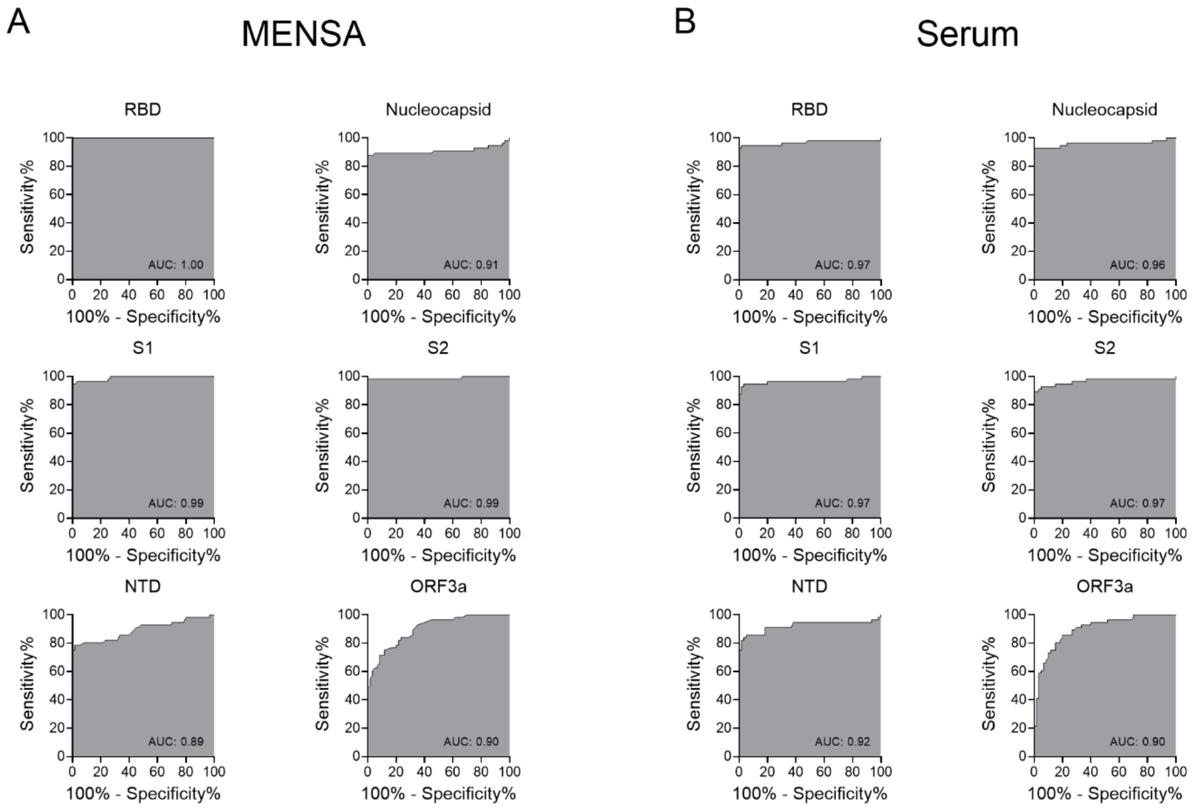


**Supplementary Figure 2. MENSA anti-RBD IgG offers superior diagnostic value while MENSA anti-Nucleocapsid IgG offers a non-spike option for natural infection.** Receiver operating characteristic (ROC) curves were performed for anti-SARS-CoV-2 IgG levels in MENSA (A) and serum (B) samples collected from healthy adults with no SARS-CoV-2 exposure and acute infected primary wild type patients. The Area Under the Curve (AUC) measurement provides an index of diagnostic potential. An AUC near 0.5 suggests no diagnostic value while an AUC near 1.0 indicates strong diagnostic potential.

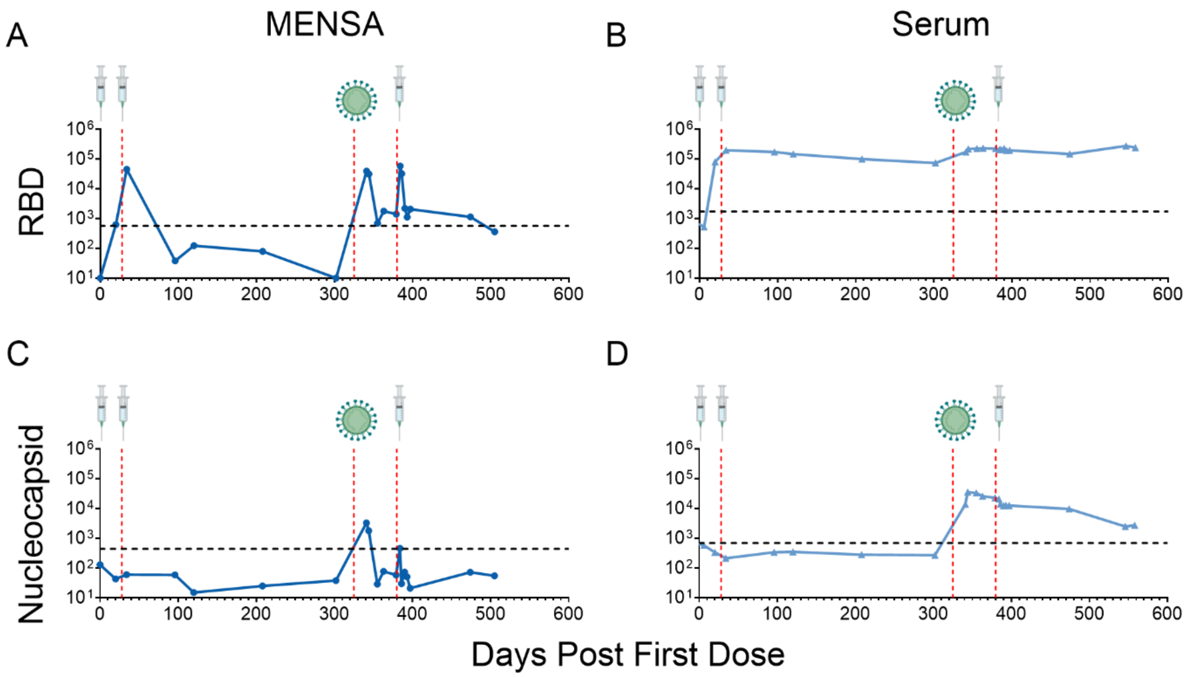


**Supplementary Figure 3. Kinetics of MENSA and serum after three vaccine doses and a breakthrough infection.** Line graphs show MENSA (dark blue) and serum (light blue) IgG antibody responses to S1-RBD (A,B) and Nucleocapsid (C,D) over time for a single patient through two primary Moderna COVID-19 mRNA vaccine doses in February-March 2021, a breakthrough Omicron infection in December 2021, and a third booster dose of mRNA COVID-19 vaccine in February 2022. All values are reported as average Net MFI (Median Fluorescent Intensity – Background). Red vertical dashed lines represent a new exposure event. Each vaccination dose event is symbolized as a syringe. The breakthrough infection event is symbolized as a virion. Horizontal dashed black lines represent the $C_0$ threshold of positivity for each sample and antigen combination as determined from Figure 1.

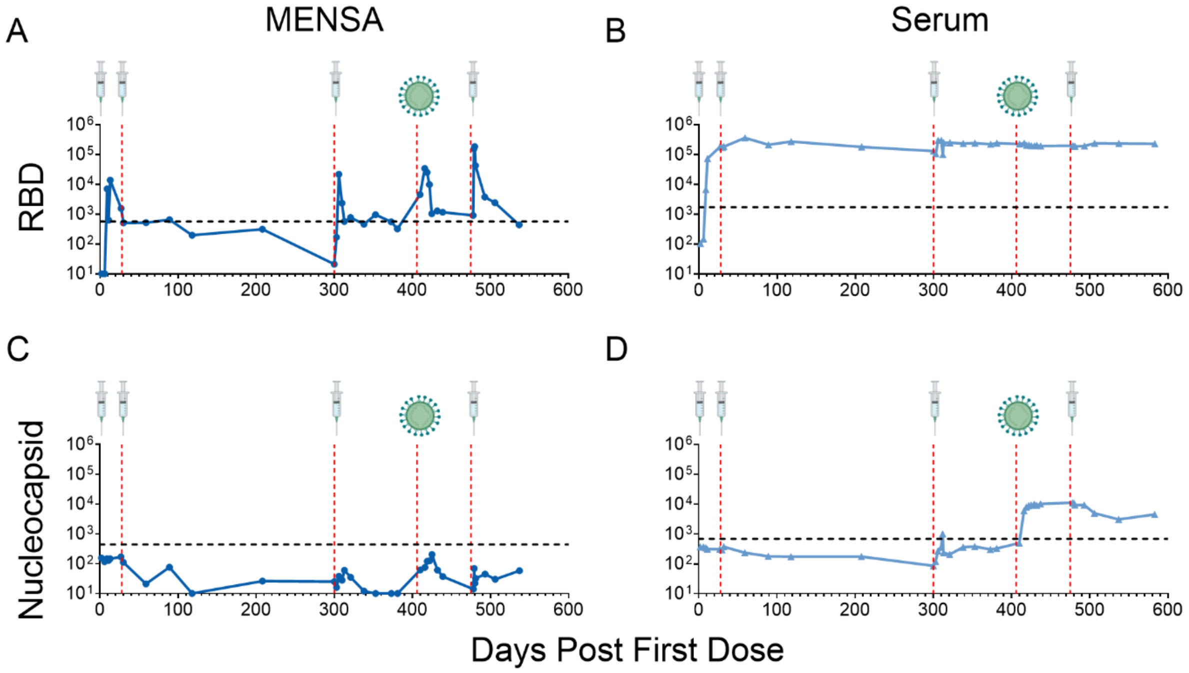


**Supplementary Figure 4. Kinetics of MENSA and serum after four vaccine doses and a breakthrough infection.** Line graphs show MENSA (dark blue) and serum (light blue) IgG antibody responses to S1-RBD (A,B) and Nucleocapsid (C,D) over time for a single patient through two primary Moderna COVID-19 mRNA vaccine doses in January 2021, a third booster dose of mRNA COVID-19 vaccine in October 2021, a breakthrough Omicron infection in February 2022, and a fourth mRNA COVID-19 vaccine dose in April 2022. All values are reported as average Net MFI (Median Fluorescent Intensity – Background). Red vertical dashed lines represent a new exposure event. Each vaccination dose event is symbolized as a syringe. The breakthrough infection event is symbolized as a virion. Horizontal dashed black lines represent the $C_0$ threshold of positivity for each sample and antigen combination as determined from Figure 1.
